# Prevalence of RT-qPCR-detected SARS-CoV-2 infection at schools: First results from the Austrian School-SARS-CoV-2 prospective cohort study

**DOI:** 10.1101/2021.01.05.20248952

**Authors:** Peter Willeit, Robert Krause, Bernd Lamprecht, Andrea Berghold, Buck Hanson, Evelyn Stelzl, Heribert Stoiber, Johannes Zuber, Robert Heinen, Alwin Köhler, David Bernhard, Wegene Borena, Christian Doppler, Dorothee von Laer, Hannes Schmidt, Johannes Pröll, Ivo Steinmetz, Michael Wagner

## Abstract

**Background:** The role of schools in the SARS-CoV-2 pandemic is much debated. We aimed to quantify reliably the prevalence of SARS-CoV-2 infections at schools detected with reverse-transcription quantitative polymerase-chain-reaction (RT-qPCR).

**Methods:** This nationwide prospective cohort study monitors a representative sample of pupils (grade 1-8) and teachers at Austrian schools throughout the school year 2020/2021. We repeatedly test participants for SARS-CoV-2 infection using a gargling solution and RT-qPCR. We herein report on the first two rounds of examinations. We used mixed-effect logistic regression to estimate odds ratios and robust 95% confidence intervals (95% CI).

**Findings:** We analysed data on 10734 participants from 245 schools (9465 pupils, 1269 teachers). Prevalence of SARS-CoV-2 infection increased from 0.39% at round 1 (95% CI 0.28-0·55%, 29 September-22 October 2020) to 1·39% at round 2 (95% CI 1·04-1·85%, 10-16 November). Odds ratios for SARS-CoV-2 infection were 2·26 (95% CI 1·25-4·12, P=0·007) in regions with >500 vs. ≤500 inhabitants/km^2^, 1·67 (95% CI 1·42-1·97, P<0·001) per two-fold higher regional 7-day incidence, and 2·78 (95% CI 1·73-4·48, P<0·001) in pupils at schools with high/very high vs. low/moderate social deprivation. Associations of community incidence and social deprivation persisted in a multivariable adjusted model. Prevalence did not differ by average number of pupils per class nor between age groups, sexes, pupils vs. teachers, or primary (grade 1-4) vs. secondary schools (grade 5-8).

**Interpretation:** This monitoring study in Austrian schools revealed SARS-CoV-2 infection in 0·39%-1·39% of participants and identified associations of regional community incidence and social deprivation with higher prevalence.

**Funding:** BMBWF Austria.

## Introduction

The SARS-CoV-2 pandemic poses unprecedented challenges on our educational systems.^1^ As part of wider strategies to contain the spread of the SARS-CoV-2 virus, many countries have devised measures at schools with the aim of reducing infection risk. These measures include adapted in-person learning (e.g. reduced class sizes, staggered time tables, wearing of masks), complete school closures coupled with virtual learning, or hybrid models.^2^ School closures represent a very effective non-pharmaceutical intervention to reduce the transmission of SARS-CoV-2,^3^ but have many adverse consequences.^1^ Thus, there is extensive debate about the role of schools and children in the SARS-CoV-2 pandemic.^1,4^

Several prior studies have examined representative samples of the general population to assess how frequently SARS-CoV-2 infections occur in children compared to adults. For instance, seroepidemiological studies^5–7^ showed a slightly lower prevalence of SARS-CoV-2 antibodies in children than in adults, but it remains unclear whether this difference is due to reduced exposure associated with schools closures, a distinct immune response, or – indeed – reduced susceptibility. Screening studies of the general population in the UK demonstrated an increase in SARS-CoV-2 prevalence in children from mid-September to December 2020 when schools were open, and a reduction in prevalence in January and February 2021 upon school closure in response to the B.1.1.7 variant of concern.^8–10^ In contrast to these studies in the general population, evidence from studies with a specific focus on schools is sparse and is largely restricted to contact-tracing and notification-based studies,^11^ rather than large-scale screening of schools for SARS-CoV-2 infection. As children are more likely to be asymptomatic than adults after a SARS-CoV-2 infection,^1^ large-scale screening studies are particularly needed for comprehensively investigating their role in the SARS-CoV-2 pandemic.

We therefore designed the School-SARS-CoV-2 Study, a nationwide prospective cohort study that screens pupils and teachers at schools in Austria for the occurrence of SARS-CoV-2 infection. We planned to repeatedly examine study participants repeatedly in 3-5 week intervals during the school year 2020/2021. In the present analysis, we analysed data from the first two rounds of examinations with two main aims: (i) to reliably quantify the prevalence of SARS-CoV-2 infection detected with reverse-transcription quantitative polymerase-chain-reaction (RT-qPCR) and (ii) to determine factors that may be associated with a higher or lower prevalence, thereby informing upcoming public health policies.

## Methods

### Study population

The School-SARS-CoV-2 Study is a nationwide prospective cohort study that monitors a representative sample of pupils and teachers in Austrian schools for presence of RT-qPCR-detected SARS-CoV-2 infection. Throughout the school year 2020/2021, repeat measurements are conducted every 3-5 weeks during periods that are not affected by school closures. After excluding schools with less than 20 pupils from the sampling frame, we randomly selected a total of 250 schools to participate in the study, corresponding to 5·6% of all schools in Austria. The selection process was stratified by federal states, employed selection probabilities proportional to the numbers of pupils enrolled at the schools, and involved primary schools (grade 1-4) and secondary schools (grade 5-8). In the current paper, we report on the first two rounds of examinations conducted between 28 September and 22 October 2020 (round 1) and between 10 and 16 November (round 2). At the time of these examinations, schools conducted face-to-face teaching with the following mitigation measures in place: (i) reduction of contacts within schools (i.e. avoidance of contact between pupils in different classes, with only teachers switching between student cohorts), (ii) physical distancing ≥1 meter in communal areas, (iii) classroom ventilation by opening windows hourly, (iv) wearing of masks in communal areas if physical distancing could not be respected (round 1) or at all times in communal areas (round 2), and (v) sports classes with physical distancing ≥2 meter (indoors or outdoors during round 1, outdoors whenever possible during round 2). Indoor music lessons and singing as well as school events could take place during round 1, but were not allowed during round 2. If required due to the regional epidemiological situation (e.g. high community incidence, high number of cases at school, uncertain source of infection), additional mitigation measures targeted to the affected schools beyond the aforementioned measures were activated.

Within each school, 60 pupils spread across all classes were invited randomly to participate in the study. In small schools with a total number of pupils less than 60, all pupils at the respective school were invited. The study population was supplemented with a random selection of teachers at a target sampling proportion of 1:10, compared to the number of pupils selected at a school. To maximise participation rates, invitation letters included extensive information material about study aims, benefits and potential harms and consent forms in five different languages (German, English, Turkish, Romanian and Bosnian-Croatian-Serbian). We also created publicly available videos^12^ in the same five languages to explain the process of the gargling test.

The study received ethics approvals by the ethics committees of the Medical University of Graz (no. 32-672 ex 19/20), Medical University of Innsbruck (no. 1319/2020), the Johannes Kepler University of Linz (no. 1222/2020), and the University of Vienna (no. 00591/2020). Written informed consent was obtained from (i) teachers, (ii) participants and their legal representative for pupils aged 14 years or older, or (iii) their legal representative only for pupils younger than 14 years, according to the ethical approval.

### SARS-CoV-2 detection by RT-qPCR

To test participants for presence of RT-qPCR-detected SARS-CoV-2 infection, they were asked to gargle 5 ml of a physiological saline solution (0·9%) or a modified Hank’s balanced salt solution (CaCl_2_ x 2H_2_O 1·26 mmol, MgCl_2_ x 6H_2_O 0·493mM, MgSO_4_ x 7H_2_O 0·41 mM, KCl 5·33 mM, KH_2_PO_4_ 0·44 mM, NaHCO3 4·17 mM, NaCl 137·93 mM, Na_2_HPO_4_ 0·34 mM, D-Glucose 5·56 mM) for a total of 60 seconds. Participants were asked not to eat or drink for at least one hour before gargling. After gargling, the specimen was first transferred to a 50 ml falcon tube and subsequently pipetted into a sample tube, cooled at a temperature of 2-8 °C, and transported cooled by courier to one of the four study laboratories for further analysis within a day. To assure that gargling specimen was collected in a standardised manner, school doctors, their assistants, and participants received access to training videos and printed material with detailed step-by-step instructions. Gargling for sample generation is part of the Austrian test strategy outlined by the Austrian Ministry of Health and is widely applied in Austria also for diagnostic testing. Gargling has been demonstrated to produce comparable sample quality like throat swab samples for other respiratory viruses^13^ and has also been applied successfully for the detection of SARS-CoV-2.^14–17^

At the laboratories, sample inactivation, RNA extraction, and RT-qPCR detection of SARS-CoV-2 was performed according to previously established protocols (for details, see Supplementary Material). Gargling samples were analysed in pools with a maximal pool size of 10. Positively tested pools were opened and samples were analysed individually. For all positively reported samples at least two viral genes were detected. SARS-CoV-2 RT-qPCR results were immediately reported to the study participants and school administrations via text messaging and/or email. Whenever a positive test result was obtained, the local health authorities were also informed instantaneously according to Austrian law.

### Additional participant and school characteristics available in our study

In addition to the exact time point and the SARS-CoV-2 RT-qPCR test result, data were recorded on the participants’ age and sex and – for pupils – the grade and class they currently attend. At the school level, we collated information on (i) the type of school (primary vs. secondary school), (ii) the geographical location, (iii) the total number of teachers, pupils, and classes at the school, (iv) population density at the municipality in which the school is located, and (v) an average social deprivation index for the pupils attending the school.

The social deprivation index had been ascertained in 2013 using methods described previously^18^. In brief, it combined information on four distinct domains: (i) highest level of education of the pupil’s parents, (ii) current occupation of the pupil’s parents, (iii) migration background, defined as both parents born in a foreign country (OECD definition), and (iv) first language other than German. Following the recommendation of Bruneforth *et al*.,^19^ we categorised the social deprivation index score, as “low” (score 100-<115), “moderate” (score 115-<125), “high” (score 125-135) and “very high” (score >135). Finally, we obtained data on the regional 7-day incidence of documented COVID-19 cases for all 94 districts in Austria via the publicly available data and dashboard of the Austrian health authority^20^ and merged these data with the school datasets according to districts and time points of the gargling tests.

### Statistical methods

Findings herein are reported according to the STROBE statement (**Supplementary Table 1**). In planning the sample size for this study, we estimated the widths of 95% confidence intervals of SARS-CoV-2 infection prevalences ranging between 0 and 1%, assuming a design effect of 4·0, and considered a sample size involving 11900 pupils and 1200 teachers recruited across 250 schools to afford sufficient statistical power. For instance, based on the assumed design effect, we expected prevalences of 0·15%, 0·40%, and 0·70% to be associated with 95% confidence intervals of 0·05-0·36%, 0·21-0·68%, and 0·45-1·05%, respectively.

In descriptive analyses, we summarised categorical variables as counts and percentages and continuous variables as means and standard deviations (if approximately normally distributed). We tested whether characteristics of participating schools were associated with each other using χ^2^-tests. In analyses of the prevalence of RT-qPCR-detected SARS-CoV-2 infection (ie, overall prevalence, differences in the prevalence across the two rounds of examinations, and differences in the prevalence across subgroups), to take into account the clustering of the data at the school level, we calculated 95% confidence intervals from robust standard errors based on clustered Sandwich estimators^21,22^. Participants with a positive test result at round 1 were censored at round 2, as they are not considered to be at risk after having already experienced the infection. Odds ratios for RT-qPCR-detected SARS-CoV-2 infection were estimated using mixed-effect logistic regression models with random intercepts at the participant level. In a first step, we estimated unadjusted odds ratios for participant and school characteristics available in our study (listed above) using univariable models. For characteristics significantly associated with SARS-CoV-2 infection in the univariable model, we then estimated multivariable adjusted odds ratios to control for potential confounding. The multivariable adjusted model included the following explanatory variables: type of school (secondary vs. primary), local population density (>500 vs. ≤500 inhabitants/km^2^), average number of pupils per class (entered as linear term), type of participant (teachers vs. pupils), log-transformed regional 7-day incidence (entered as linear term), and the social deprivation index (high/very high vs. low/moderate). Furthermore, we conducted post-hoc subgroup analyses that compared the strengths of associations in teachers vs. pupils using formal tests for interaction. We conducted analyses with Stata version 14·1 MP. We used two-sided statistical tests and considered P values ≤0·05 as statistically significant.

## Results

### Study population

The flow chart of the schools and participants involved in our study is shown in **Figure 1**. The first round of examinations was conducted between 29 September 2020 and 22 October 2020 and covered 243 out of the 250 eligible schools (97·2%) across all nine federal states of Austria. Round 1 included 10464 individuals, of which 10156 (97·1%) had a valid gargle test. The median regional 7-day incidence of SARS-CoV-2 cases in the general population at the time of the examinations was 75 per 100,000 inhabitants (IQR 43-125). The second round of examinations was conducted between 10 and 16 November 2020, ending early due to the closure of schools as part of a wider lockdown in Austria employed on 17 November 2020. Round 2 involved 88 schools across five federal states of Austria (i.e. Burgenland, Lower Austria, Upper Austria, Vorarlberg, Vienna). Of 3785 individuals that took part in round 2, 3745 had a valid gargle test. The corresponding median regional 7-day incidence in the general population was 419 per 100,000 inhabitants (IQR 392-641). Overall, across the two rounds of examinations, data from 245 schools, 10734 participants, and 13901 measurements were available for further analysis.

**Figure 1.**
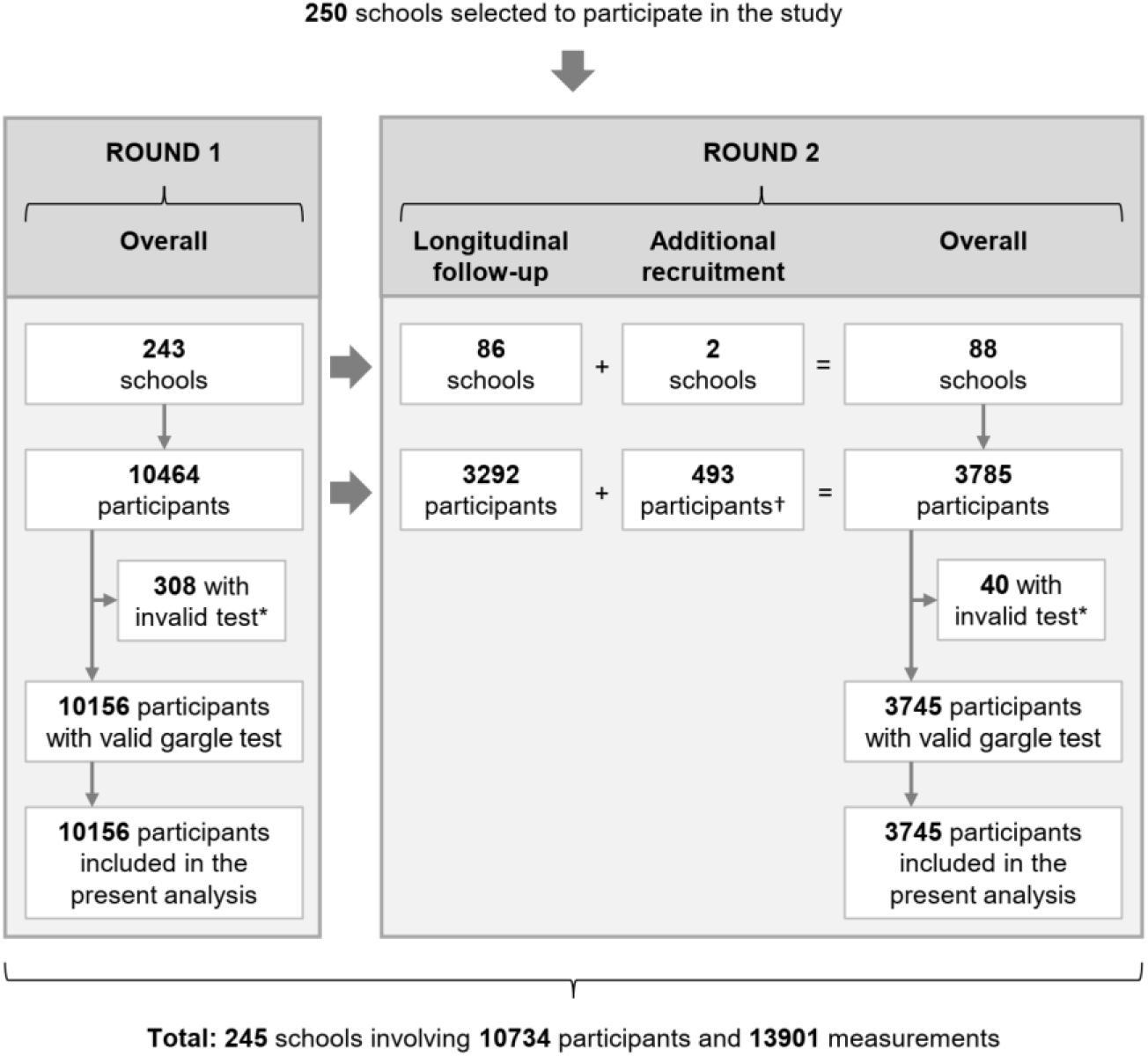
Flow-chart of the recruitment into the study. *Mostly due to transfer of insufficient volumes to the test tubes by the school doctors. †The 493 participants additionally recruited at round 2 are composed of 123 individuals recruited at the two additional schools participating in round 2 (111 pupils, 12 teachers) and 370 individuals recruited at schools that already participated in round 1.

Key characteristics of schools and participants are provided in **Table 1** and **Supplementary Figure 1**. The number of participants was distributed equally across primary and secondary schools (**Table 1**). At the schools participating in the study, the median total number of pupils was 230 (interquartile range [IQR] 147-331), the median total number of teachers was 27 (IQR 17-43), and classes consisted – on average – of 21 pupils (IQR 18-23) (**Supplementary Figure 1**). Schools each recruited a median of 40 pupils (IQR 29-50) and 6 teachers (IQR 5-6) into our study. On average, teachers constituted 11.8% of a school’s study sample (IQR 10·0-14·6%).

**Table 1.** Descriptive summary of schools and participants investigated as part of the School-SARS-CoV-2 Study.

|  | No. of schools (%) | No. of participants (%) |
| --- | --- | --- |
| **Total** | 245 (100.0%) | 10734 (100.0%) |
| **Type of school** |  |  |
| Primary school | 129 (52.7%) | 5367 (50.0%) |
| Secondary school | 116 (47.3%) | 5367 (50.0%) |
| **Geographical location by federal state\*** |  |  |
| Burgenland | 15 (6.1%) | 669 (6.2%) |
| Carinthia | 15 (6.1%) | 665 (6.2%) |
| Lower Austria | 46 (18.8%) | 2072 (19.3%) |
| Upper Austria | 42 (17.1%) | 1716 (16.0%) |
| Salzburg | 15 (6.1%) | 610 (5.7%) |
| Styria | 29 (11.8%) | 1287 (12.0%) |
| Tyrol | 18 (7.3%) | 583 (5.4%) |
| Vorarlberg | 15 (6.1%) | 579 (5.4%) |
| Vienna | 50 (20.4%) | 2553 (23.8%) |
| **Local population density** |  |  |
| ≤100 inhabitants/km<sup>2</sup> | 23 (9.4%) | 893 (8.3%) |
| >100-250 inhabitants/km<sup>2</sup> | 47 (19.2%) | 1863 (17.4%) |
| >250-500 inhabitants/km<sup>2</sup> | 34 (13.9%) | 1396 (13.0%) |
| >500-10000 inhabitants/km<sup>2</sup> | 126 (51.4%) | 5781 (53.9%) |
| >10000 inhabitants/km<sup>2</sup> | 15 (6.1%) | 801 (7.5%) |
| **Social deprivation index†** |  |  |
| Low/moderate | 183 (75.6%) | 7833 (73.9%) |
| High/very high | 59 (24.4%) | 2762 (26.1%) |
| **Average no. of pupils/class** |  |  |
| ≤20 pupils/class | 104 (42.4%) | 4087 (38.1%) |
| >20 pupils/class | 141 (57.6%) | 6647 (61.9%) |

\*In comparison, the overall number of pupils per federal state were 21056 (3.1%) in Burgenland, 41322 (6.1%) in Carinthia, 128962 (19.0%) in Lower Austria, 120603 (17.8%) in Upper Austria, 42863 (6.3%) in Salzburg, 89844 (13.2%) in Styria, 57570 (8.5%) in Tyrol, 33285 (4.9%) in Vorarlberg, and 143617 (21.1%) in Vienna. †Information on the social deprivation index was not available for three schools involving 139 participants.

**Supplementary Table 2** illustrates pairwise associations between available school characteristics. Higher local population density was associated with a greater social deprivation index (P<0·001) and higher average number of pupils per class (P<0·001). As expected, primary schools had smaller class sizes than secondary schools (P<0·001).

### Prevalence of RT-qPCR-detected SARS-CoV-2 infection

**Table 2** presents prevalence of RT-qPCR-detected SARS-CoV-2 infection at round 1 and 2 of our study. At round 1, 40 out of 10156 participants were tested positive, corresponding to a prevalence of 0·39% (95% confidence interval [CI]: 0·28-0·55%). Prevalence was 0·37% among pupils (95% CI 0·26-0·53, 33 out of 8934) and 0·57% among teachers (95% CI 0·25-1·32, 7 out of 1222).

**Table 2.** Participant characteristics and prevalence of RT-qPCR-detected SARS-CoV-2 infection at the two rounds of examinations conducted between 28 September and 16 November 2020 within the School-SARS-CoV-2 Study.

|  | Round 1 | Round 2 | Overall |
| --- | --- | --- | --- |
| **Assessment of participants** |  |  |  |
| No. of schools | 243 | 88 | 245 |
| Federal states involved in assessment | All | Burgenland,<br>Lower Austria,<br>Upper Austria,<br>Vorarlberg,<br>Vienna | All |
| Median date of assessment in the year 2020 (range) | 12.10.<br>(28.09.-22.10.) | 11.11.<br>(10.11.-16.11.) | 14.10.<br>(28.09.-16.11.) |
| Regional 7-day incidence in the general population (per 100,000 inhabitants), median (IQR) | 75 (43-125) | 419 (392-641) | 114 (53-357) |
| **No. of participants** | 10156 | 3745 | 10734 |
| **Participants by type of school** |  |  |  |
| No. at primary school (%) | 5029 (50%) | 2046 (55%) | 5367 (50%) |
| No. at secondary school (%) | 5127 (50%) | 1699 (45%) | 5367 (50%) |
| **Teachers** |  |  |  |
| No. of teachers (%) | 1222 (12%) | 450 (12%) | 1269 (12%) |
| Age (years), mean (SD) | 43.9 (11.8) | 44.3 (11.9) | 44.0 (11.8) |
| No. of females (%) | 986 (81%) | 372 (83%) | 1026 (81%) |
| **Pupils** |  |  |  |
| No. of pupils (%) | 8934 (88%) | 3295 (88%) | 9465 (88%) |
| Age (years), mean (SD) | 9.9 (2.4) | 9.8 (2.4) | 9.8 (2.4) |
| No. of females (%) | 4291 (48%) | 1592 (48%) | 4541 (48%) |
| **RT-qPCR-detected SARS-CoV-2 infection** |  |  |  |
| No. of cases | 40 | 52 | 92 |
| Period prevalence (95% CI) | 0.39%<br>(0.28-0.55%) | 1.39%<br>(1.04-1.85%) | 0.86%<br>(0.67-1.10%) |

CI denotes confidence interval, SD denotes standard deviation. 95% CI were calculated from robust standard errors estimated based on clustered Sandwich estimators.

At round 2, 52 additional participants out of 3745 participants were tested positive. This corresponded to a prevalence of 1·39% (95% CI 1·04-1·85%) and was significantly higher than at round 1. The odds ratio (OR) for SARS-CoV-2 infection in round 2 compared to round 1 was 3·56 (95% CI 2·32-5·46, P<0·001). This result was confirmed in sensitivity analysis restricted to the 3178 individuals that had valid gargling test results at both rounds (OR 4·44, 95% CI 1·48-13·32, P<0·001). Prevalence was 1·52% among pupils (95% CI 1·13-2·04, 50 out of 3295) and 0·44% among teachers (95% CI 0·11-1·79, 2 out of 450).

### Association of participant and school characteristics with SARS-CoV-2 infection

In **Figure 2**, we investigated whether participant and school characteristics were associated with the odds of being tested positive for RT-qPCR-detected SARS-CoV-2 infection. In the unadjusted model (**Figure 2a**), significant positive associations were detected for local population density, regional 7-day incidence in the general population, and the social deprivation index. The ORs for RT-qPCR-detected SARS-CoV-2 infection were 2·26 for schools located in regions with >500 vs. ≤500 inhabitants per km^2^ (95% CI 1·25-4·12, P=0·007), 1·67 for a two-fold higher regional 7-day community incidence (95% CI 1·42-1·97, P<0·001), and 2·78 in pupils at schools with a high or very high social deprivation index compared to their counterparts (95% CI 1·73-4·48, P<0·001).

**Figure 2.**
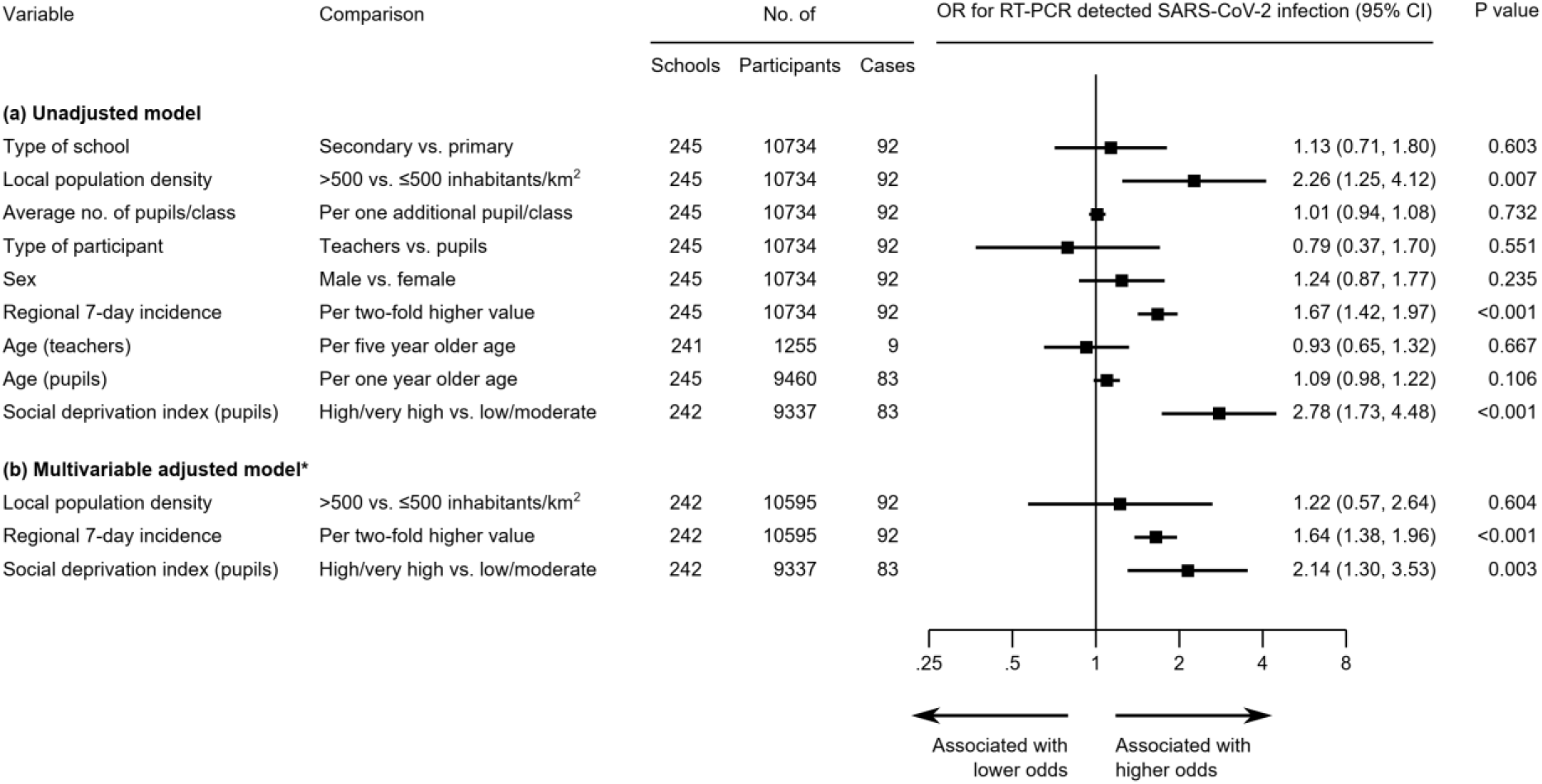
Odds ratios for RT-qPCR-detected SARS-CoV-2 infection at the two rounds of examinations according to participant and school characteristics in an unadjusted model (Panel a) and a multivariable adjusted model* (Panel b). CI denotes confidence interval, OR odds ratio, and RT-qPCR real-time polymerase chain reaction. The analysis involved data from round 1 and 2 of the School-SARS-CoV-2 Study. Odds ratios were estimated using mixed-effect logistic regression models with random intercepts at the participant level. 95% confidence intervals were calculated from robust standard errors estimated from clustered Sandwich estimators. Variables that were associated with RT-qPCR-detected SARS-CoV-2 infection at a significant level of ≤0.05 in the unadjusted model (Panel a) were included in the multivariable adjusted model (Panel b). *The multivariable adjusted model included the following explanatory variables: type of school (secondary vs. primary), local population density (>500 vs. ≤500 inhabitants/km^2^), average number of pupils per class (entered as linear term), type of participant (teachers vs. pupils), sex (male vs. female), log-transformed regional 7-day incidence (entered as linear term), and the social deprivation index (high/very high vs. low/moderate).

In the multivariable adjusted model (**Figure 2b**), only regional 7-day community incidence in the general population and the social deprivation index retained statistical significance, with ORs of 1·64 (95% CI 1·38-1·96, P<0·001) and 2·14 (95% CI 1·30-3·53, P=0·003), respectively. There was no significant association for local population density in the multivariable adjusted model (P=0·604).

In a post-hoc subgroup analyses, we investigated whether associations differed between pupils and teachers. Regional 7-day community incidence was more strongly associated with RT-qPCR-detected SARS-CoV-2 infection in pupils (OR 1·81, 95% CI 1·53-2·15) than in teachers (OR 0·97, 95% CI 0·76-1·25) (P value for interaction<0·001). There was no effect modification of the OR for age (P=0·102) or the OR for local population density (P=0·078).

### No. of cases recorded per school

At the first round of examinations, 209 schools recorded no cases of SARS-CoV-2 infection (86·0%), 28 schools recorded one case (11·5%), and 6 schools recorded two cases (2·5%). At second round, 52 schools recorded no cases (59·1%), 23 schools recorded one case (26·1%), 10 schools recorded two cases (11·4%), and 4 schools recorded three cases (3·4%). The associations of community incidence and social deprivation index with infection frequencies at schools is visualised in **Figure 3a** and **Figure 3b**.

**Figure 3.**
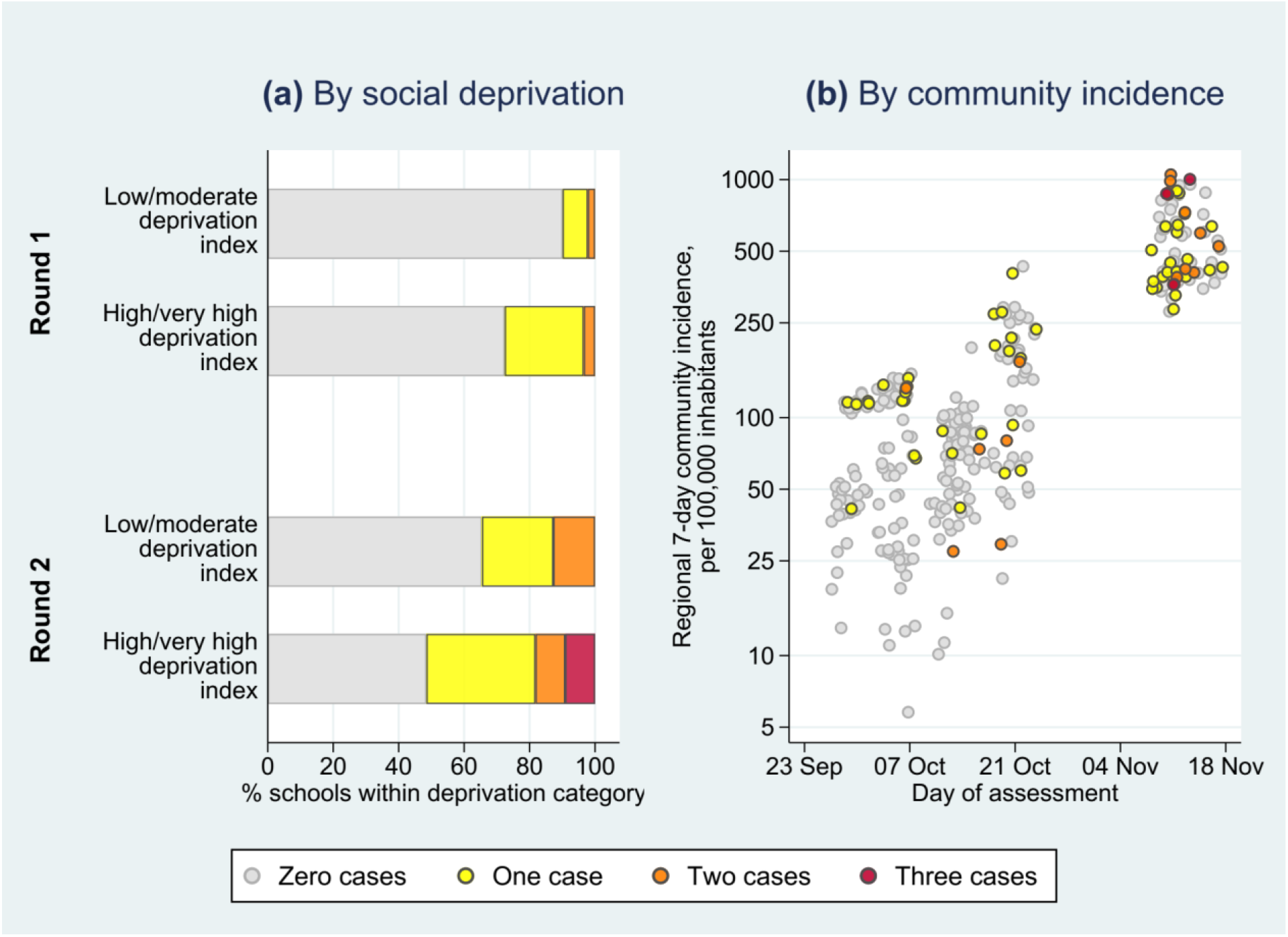
Number of RT-qPCR-detected SARS-CoV-2 cases per school according to the social deprivation index (Panel a) and regional 7-day community incidence (Panel b). The regional 7-day community incidence shown in the graph is the incidence in the general population in the district, in which the school is located, and at the time point of the gargling test, obtained from the dashboard of the Austrian health authority. The methods used to ascertain social deprivation are described in the methods section.

### C_t_ values of RT-qPCR tests of positively tested participants

**Supplementary Figure 2** shows the C_t_ values in unpooled samples of all positively tested participants for the RT-qPCR assays used in the four laboratories. There was no statistically significant difference in C_t_ values between pupils and teachers. However, C_t_ values were inversely associated with older age of the pupils for some RT-qPCR assays. As expected for a screening study in asymptomatic individuals, most C_t_ values were relatively high. 34% of the pupils and 11% of the teachers had a C_t_ value <30 in at least one of the RT-qPCR assays (P=0.263 in Fisher’s exact test) and may thus be considered to be potentially infectious at the time of sampling.

## Discussion

The present study reports on the prevalence of RT-qPCR-detected SARS-CoV-2 infection at Austrian schools involving a total of 245 schools and 10734 individuals. At the first round of examinations conducted between 28 September and 22 October 2020, we detected SARS-CoV-2 infection in 0·39% of the study participants (95% CI 0·25-0·55%). At the second round conducted between 10 and 16 November, we observed an approximately 3·6-fold higher prevalence with a point estimate of 1·39% (95% CI 1·04-1·85%). Furthermore, among a range of participant and school characteristics, regional 7-day incidence in the general population and social deprivation emerged as relevant factors associated with presence of RT-qPCR-detected SARS-CoV-2 infection at the participating schools. Collectively, the study provides crucial evidence about the extent and determinants of SARS-CoV-2 infection at schools, thereby informing decision making about in-person education at Austrian schools and elsewhere in upcoming months.

To which extent children are inflicted by SARS-CoV-2 and to which extent opening or closing of schools impacts the dynamics of the SARS-CoV-2 pandemic is much debated.^1,4^ One aspect of this debate concerns the occurrence of SARS-CoV-2 infection in children compared to adults. In our study, we detected SARS-CoV-2 infection in 1·39% (95% CI 1·04-1·85%) of study participants at the second round of examinations (10-16 November). This prevalence was somewhat less than the screen-detected prevalence of 2·12% in people aged ≥16 years, which was observed by a different nationwide population-based study^23^ conducted at a similar time frame (12-14 November, 48 out of 2263 tests positive based on nasopharyngeal swabs). A crude comparison of the two studies yields a prevalence ratio of 0·65 (95% CI 0·44-0·97, P=0·032), although one should interpret this ratio with caution due to potential selection biases (e.g. participation rate 28·9% in the latter study^23^) and different testing methods (e.g. gargling vs. nasopharyngeal swabs). Importantly, samples were not pooled in the nationwide population-based study, while, in our study, pool sizes from 3 to 10 were used. Pooling has been used for the diagnosis of influenza^24^ and has been adapted and employed successfully in the field of SARS-CoV diagnostics^25,26^. In RT-qPCR analyses, C_t_ values increase with template dilution and pooling of 10 samples would lead to a theoretically calculated C_t_ value increase of 3.3. Our school screening is thus expected to overlook positive samples with C_t_ values above 36-37 in pools with a size of 10.

This prevalence ratio is consistent with several population-based studies that investigated seroprevalence of antibodies to SARS-CoV-2 across the age spectrum.^27^ For instance, the seroepidemiological study in Spain ENE-COVID reported a continuous rise in seropositivity from the youngest age groups (1·1% <1 yr, 2·1% 1-4 yrs, 3·1% 5-9 yrs, 4·0% 10-14 yrs) into adulthood with a plateau at around 6% at 45 years of age or older.^5^ Differences by age were also observed in regional seroprevalence studies in Switzerland (SEROCoV-POP Study^6^) and Austria (Ischgl Study^7^). While these differences may stem from lower susceptibility to SARS-CoV-2 in children (e.g. due to reduced *ACE2* expression in the nasal epithelium^28,29^), they could also arise from reduced exposure (e.g. due to school closures) or a milder clinical course of the infection coupled with a distinct immune response characterised by absence of anti-nucleocapsid IgG antibodies^30^. Furthermore, in light of changing policies on school-based preventive measures and potential seasonal variation, prevalence estimates of SARS-CoV-2 infection in children may be highly volatile. For instance, the UK-based REACT-1 study^8,10^ reported that prevalence in children aged 5-17 years had increased sharply from mid-September to December 2020, surpassing the prevalence observed in adults, and had been reduced in January and February 2021 upon school closure in response to the B.1.1.7 variant of concern, with similar time trends reported by the UK Office of National Statistics^9^.

Our study has identified several factors associated with higher odds of a SARS-CoV-2 case in the setting of in-person education at schools. The strongest link was observed with regional 7-day incidence in the general population, associated with 1·66-fold odds (95% CI 1·39-1·99, P<0·001) for each doubling of community incidence. This observation is in agreement with a Public Health England report that regional community incidence was significantly associated with the risk of COVID-19 outbreaks at schools.^11^ Our study crucially extends the existing evidence to the months in autumn and to a markedly higher community incidence (median 7-day incidence 114 vs. ~6 per 100.000). Interestingly, post-hoc subgroup analyses revealed that the association of community incidence with the odds of a SARS-CoV-2 case was confined to pupils and not to teachers, which may be attributable to differential testing strategies or differential adherence to preventive measures in these two groups. Another important factor associated with higher odds of SARS-CoV-2 cases was social deprivation. While this finding highlights the need for additional carefully-targeted support of affected children, the underlying mechanisms remain to be determined. These likely go beyond school-related factors and may include less self-isolation and physical distancing,^31^ cramped living conditions,^31^ and lack of possibilities for parents to work from home, also coupled with challenges in taking care of a sick child. Of note, ecological analyses in the US^32^ and Germany^33^ have previously shown that deprived areas are affected disproportionately by SARS-CoV-2. It is also noteworthy that we detected no significant differences in SARS-CoV-2 prevalence between primary vs. secondary schools, smaller vs. larger class sizes, pupils vs. teachers, and females vs. males, nor according to the participants’ age.

Another key aspect in assessing the role of schools in the SARS-CoV-2 pandemic is the infectiousness of children. It is well established that symptomatic children have viral nucleic acids in their nasopharynx at levels comparable with adults.^26,34^ Consistently, we observed no statistically significant difference in C_t_ values of positively tested pupils and teachers in this study (**Supplementary Figure 2a**). Previous studies reported inconsistent associations between age and C_t_ value in children. While one study found significantly higher amounts of SARS-CoV-2 RNA in children younger than five years than in older children^34^, another study reported lower C_t_ values in children aged <12 years than in adults^35^. We observed for some RT-qPCR assays among pupils an increasing amount of viral RNA with increasing age (**Supplementary Figure 2b**). As no correlation of the amount of human RNA detected in a RT-qPCR control reaction with the age of the pupils was detectable (**Supplementary Figure 2b**), it appears unlikely that these differences were caused by age-related differences in executing the gargling procedure. However, it should be noted that interpretation of this results is complicated by the fact that the RT-qPCR analyses were done in four different laboratories partly using different starting volumes and protocols (although lab-to-lab comparison revealed no major differences in C_t_ value distributions between labs, see Supplementary Methods for details). Children do not only have comparable viral loads to adults, but SARS-CoV-2 viruses from children can also be cultured in vitro, suggesting that transmission from them is plausible.^36^

Actual infectiousness is investigated best in contact tracing studies conducted within schools and/or households. Some of these studies indicate that children may be somewhat less infectious than adults.^1^ Analyses based on notifications of SARS-CoV-2 cases showed low transmission rates in Ireland before school closures^37^ and in Australian schools operating at reduced physical attendance^38^, but these analyses did not include asymptomatically infected children. In Germany, after reopening of schools, health authorities identified 2·2 outbreaks per week with four cases per outbreak on average.^39^ In England, outbreak analyses largely based on symptomatic cases showed that staff members were more frequently the seeding case in schools than students.^11^ In Austria, health authorities linked 4·3% of the SARS-CoV-2 infections recorded between 5 October and 15 November 2020 with a traceable source of infection (42%) to educational settings^40^, but it should be kept in mind that infected children are often asymptomatic and that children as contact persons were not systematically tested. A large-scale household study in South Korea suggested infected children aged 6-11 years were less contagious than infected older children that were even more contagious than adults.^41^ It is important to stress that these reduced transmission rates have been observed against the backdrop of extensive preventive measures implemented in schools, including reduced class sizes, staggered time tables, frequent ventilation, wearing of masks, and staying home even with minimal symptoms.^2^ In contrast, a report from a US summer camp illustrated the potential efficient spread in children even if they were younger than 10 years.^42^ After a 10-day camp involving various indoor and outdoor activities, including singing and cheering, a staggering attack rate of 44% was observed among 597 participants and 51% of the children <10 years were infected. Consistent with this observation, a prospective household study from the US reported substantial transmission also when the index case was a child^43^ and in a Canadian emergency childcare centre transmission of the virus from children (including an asymptomatic person) to their parents was documented^44^.

The new SARS-CoV-2 variants of concern B.1.1.7 and B.1.351 have recently been reported in Austria.^45^ While it is unlikely that these variants already occurred in Austrian schools during the sampling periods reported in this manuscript, the RT-qPCR primers and probes applied in this study would have detected them. With the continuation of this study in March 2021 all samples positively tested will be subjected to virus genome sequencing. This analysis will help to better understand whether younger age cohorts are more affected by these variants and might also provide in schools with more cases some indications about key epidemiological parameters like number of independent virus introduction events.

The study we presented herein has several major strengths. It evaluated prevalence of SARS-CoV-2 detection at a time during which in-person education at school was in place nationwide (for pupils in grade 1-8). Furthermore, it involved a large-scale sample of schools across Austria, thereby enhancing generalisability of our findings. Our study also has several limitations. First, prevalence of SARS-CoV-2 infections detected in this study likely underestimate the true burden, since symptomatic individuals (pupils and teachers) or those retained in quarantine were not present at the time of testing. Second, participation in our study was voluntary and we cannot rule out selection bias from differential uptake of study invitations. However, to limit the scope of this bias, we employed several strategies to maximise participation rates, including provision of explanatory videos and information material in several languages. Third, our study was designed to reliably quantify SARS-CoV-2 prevalence in schools. It does however not allow for evaluation of potential secondary transmission within classrooms, which would require school-based contact tracing data. Finally, the second repeat assessment of study participants was incomplete due to the decision of the Austrian government to close schools for in-person learning on 17 November 2020. We anticipate a restart of repeated screening of the cohort in early March and will continue assessments until the end of June.

In conclusion, in a large-scale study involving 245 schools in Austria, prevalence of RT-qPCR detected SARS-CoV-2 increased from 0·39% to 1·39% within a period of one month. Higher community incidence (quantified as the 7-day incidence in the school district) and social deprivation were associated with higher odds of SARS-CoV-2 at the schools. By determining SARS-CoV-2 prevalence and identifying potential contributing factors, our study provides evidence relevant to the decision making about in-person education at Austrian schools and elsewhere.

## Data Availability

Participant-level data generated and analysed during the current study are not publicly available.

## Contributors

PW, BL, RK, and MW designed the study. RK, JZ, AK, DvL, HS, IS and MW planned the logistics and/or laboratory measurements for the study. BH, ES, HS, RH, DB, WB, CD, and JP performed laboratory measurements. PW and AB performed the statistical analysis. PW, BL, RK, and MW drafted the manuscript. All authors critically revised the manuscript and agreed to be accountable for all aspects of the work.

## Acknowledgements

This study was funded by the Federal Ministry of Education, Science and Research of the Republic of Austria. We are indebted to the study participants as well as the many individuals involved in study management, participant examination, logistics, and sample analysis that made this study possible. We thank Johanna Trupke and Daniele Soroldoni from the Vienna BioCenter Core Facilities for molecular biology analytical support. We are grateful to Armin Haba, Lukas Zaminer, and Nicole Perner at the ‘Federal Institute of Quality Control of the Austrian School System’ (IQS) for their work on the sample selection for this study.

