## Supplementary Material for "Prevalence of RT-qPCR-detected SARS-CoV-2 infection at schools: First results from the Austrian School-SARS-CoV-2 prospective cohort study"

#### Supplementary methods

##### General considerations with regard to sample processing

Sample processing was performed at 4 separate laboratories across Austria. Not all laboratories were equipped with the exact same set of instruments. Laboratories at University of Vienna (1$^{st}$ and 2$^{nd}$ round), Medical University of Innsbruck (joined in 2$^{nd}$ round), Johannes Kepler University of Linz (joined in 2$^{nd}$ round) used a standard laboratory approach as detailed in **Sections 1-3**. The *in vitro* diagnostics approach for SARS-CoV-2-detection in gargling samples performed by the Medical University of Graz (1$^{st}$ round) and standard laboratory approach (2$^{nd}$ round) are detailed in **Sections 4-5**, respectively. Overall, we aimed at streamlining the individual workflows as much as possible and chose overlapping primer combinations to allow for cross-laboratory data usage (see **Supplementary Figure 3** and **Supplementary Table 3**).

##### 1. Sample inactivation and RNA extraction

Gargle samples were received in 1 ml FluidX tubes with externally threaded caps (Brooks Life Sciences, Chelmsford, MA) that were organized into 96-well racks. Several spots were left empty for RNA extraction and real-time quantitative polymerase chain reaction (RT-qPCR) controls on each rack. The outside surface of sample tubes was disinfected by immersion in 70% ethanol and racks were centrifuged at 500 × *g* for 1 min at room temperature (RT) to remove residual ethanol. Freshly prepared 2 M 1,4-dithiothreitol (DTT, 5-35 mM final conc., see **Supplementary Table 3**) was added to each tube and mixed via pipetting to reduce viscosity of the gargling specimen. Tubes were incubated for 10 min at room temperature, followed by centrifugation at 500 × *g* for 1 min at RT to enrich cellular material. Afterwards, surplus sample material was removed until 200 µl of the cell enriched sample fraction remained in each sample tube. The remaining sample material was gently mixed by pipetting up and down.

For pooling, aliquots of 50 µl of individual samples on a rack were transferred to a 96 V-bottom deep-well plate. Additional samples were added as 50 µl aliquots. Pools sizes up to 10 were either mixed by pipetting or homogenized using mechanical mixing in a KingFisher Flex (ThermoFisher Scientific, Waltham, MA). Afterwards, 100 µl of the mixed pool were transferred to a 96 V-bottom deep-well plate containing 350 µl of lysis buffer (80 mM Tris pH 6.4, 5.7 M GITC, 35 mM EDTA, 2% Triton X-100, 56 mM DTT freshly added) and incubated for 10 min at room temperature. At this step, 10 µl of RNA that had previously been extracted from positively tested samples were added in duplicate as sample processing controls to test for RNA degradation and PCR inhibition (note: this step was not performed in the laboratories at Johannes Kepler University of Linz). Automated RNA extraction with DNAse treatment was performed using the KingFisher Flex Magnetic Particle Processor System (ThermoFisher Scientific, Waltham, MA) or the CyBio Felix System (Analytik Jena, Jena, Germany) based on carboxylated magnetic bead separation[1].

**2. RT-qPCR assay**

RT-qPCR was performed with 5-7 µl of extracted RNA using the Luna Universal Probe One-Step RT-qPCR Kit with the Luna WarmStart RT Enzyme Mix (New England Biolabs, Ipswich, MA) or the BioRad OnestepRT qPCR Kit according to the manufacturers protocol in a final reaction volume of 20 µl. RNA extracts from pooled samples were analyzed using combinations of the same primers as detailed in **Supplementary Table 3** and **Supplementary Table 4**. RT-qPCR reaction controls included: (i) an RNA extract from a previously tested positive sample with a target Ct value of 25 or Synthetic SARS-CoV-2 RNA Control (1×10E4 molecules per reaction; Twist Biosciences, San Francisco, CA); (ii) an RNA extract from a previously tested negative samples; (iii) a no template control. RT-qPCR reactions were run on a CFX96 Touch Real-Time PCR Detection System (Bio-Rad, Hercules, CA) with cycling conditions of 55°C for 10 min, 95°C for 1 min and 45 cycles of 95°C for 10 s, and 55°C for 30 s. Fluorescence signal was recorded at the 55°C annealing and elongation step of each cycle. Results were analysed and Ct-values were calculated using the CFX Maestro software (Bio-Rad, Hercules, CA).

**3. RNA extraction and RT-qPCR of samples in positive pools**

If a pool was found positive, the individual samples were subjected to RNA extraction by using 100 µl of remaining DTT-treated original gargle sample as described above. RNA extracts of individual (unpooled) samples were analysed via RT-qPCR as detailed in **Supplementary Table 3** using the PCR conditions described above.

**4. In vitro diagnostics approach (Graz, used in 1$^{st}$ round)**

For the first round of the school study, presence of SARS-CoV-2 RNA of samples sent to the Molecular Diagnostics Laboratory, Diagnostic & Research Institute of Hygiene, Microbiology and Environmental Medicine at the Medical University of Graz was determined by RT-qPCR using the in vitro diagnostics/Conformité Européenne (IVD/CE)-labeled cobas SARS-CoV-2 test (Roche Molecular Systems, Branchburg, NJ, USA) for use on the cobas 6800/8800 system (Roche Molecular Diagnostics, Rotkreuz, Switzerland) according to the manufacturer's instructions.[2,3] Briefly, after disinfection of the outer surface of sample tubes in 70% ethanol and centrifugation, 10µl of freshly prepared 1 M 1,4-dithiothreitol (DTT, 5-10 mM final conc.) were added to each tube and mixed by pipetting up and down. After incubation for 10 minutes and centrifugation, an aliquot of 90 µl of 10 samples each were transferred to a new 1 ml tubes to receive pools of 10. Pooled samples were mixed by pipetting up and down and aliquots of 400 µl were added to 400 µl cobas PCR Media and tested with the SARS-CoV-2 test on the fully automated cobas® 6800/8800 system (**Supplementary Table 3**). The system uses 400 µl of the sample/media mix for extraction. When a pool was found positive, 400 µl of each individual sample were added to cobas PCR Media and tested in the same way. An RNA Internal Control, used to monitor the entire preparation and PCR amplification process, was introduced into each specimen during sample processing.

#### 5. Extraction and RT-qPCR (Graz, used in 2nd round)

For the second round of the school study, a test system consisting of extraction on the KingFisher Flex instrument using the MagMaX Viral/Pathogen Nucleic Acid Isolation Kit (Life Technologies Corporation, Austin, TX, USA) was used according to the manufacturer's instructions. Amplification was performed on the CFX96 Touch Real-time PCR Detection System for detection of SARS-CoV-2 RNA according to the protocol described above (**Section 2**, **Supplementary Table 3**). Disinfection, addition of DTT and sample pooling was performed in the same way as during the first survey (**Section 4**).

### Supplementary Tables

**Supplementary Table 1.** STROBE Checklist for Cohort Studies.

| | Item No | Recommendation | Page |
|---|---|---|---|
| **Title and abstract** | 1 | (a) Indicate the study's design with a commonly used term in the title or the abstract | 1 |
| | | (b) Provide in the abstract an informative and balanced summary of what was done and what was found | 2 |
| **Introduction** | | | |
| Background/rationale | 2 | Explain the scientific background and rationale for the investigation being reported | 5 |
| Objectives | 3 | State specific objectives, including any prespecified hypotheses | 5 |
| **Methods** | | | |
| Study design | 4 | Present key elements of study design early in the paper | 5 |
| Setting | 5 | Describe the setting, locations, and relevant dates, including periods of recruitment, exposure, follow-up, and data collection | 5-6 |
| Participants | 6 | (a) Give the eligibility criteria, and the sources and methods of selection of participants. Describe methods of follow-up | 5-6 |
| | | (b) For matched studies, give matching criteria and number of exposed and unexposed | Not applicable |
| Variables | 7 | Clearly define all outcomes, exposures, predictors, potential confounders, and effect modifiers. Give diagnostic criteria, if applicable | 6-7 |
| Data sources/ measurement | 8* | For each variable of interest, give sources of data and details of methods of assessment (measurement). Describe comparability of assessment methods if there is more than one group | 6-7 |
| Bias | 9 | Describe any efforts to address potential sources of bias | 5-7 |
| Study size | 10 | Explain how the study size was arrived at | 7-8 |
| Quantitative variables | 11 | Explain how quantitative variables were handled in the analyses. If applicable, describe which groupings were chosen and why | 8 |
| Statistical methods | 12 | (a) Describe all statistical methods, including those used to control for confounding | 8 |
| | | (b) Describe any methods used to examine subgroups and interactions | 8 |
| | | (c) Explain how missing data were addressed | 8 |
| | | (d) If applicable, explain how loss to follow-up was addressed | 8 |
| | | (e) Describe any sensitivity analyses | 8 |
| **Results** | | | |
| Participants | 13* | (a) Report numbers of individuals at each stage of study—eg numbers potentially eligible, examined for eligibility, confirmed eligible, included in the study, completing follow-up, and analysed | 8-9, Figure 1, Supplementary Figure 1 |
| | | (b) Give reasons for non-participation at each stage | |
| | | (c) Consider use of a flow diagram | |
| Descriptive data | 14* | (a) Give characteristics of study participants (eg demographic, clinical, social) and information on exposures and potential confounders | 9-10, Table 1, Table 2, Supplementary Table 2 |

| | | | |
|---|---|---|---|
| | | (b) Indicate number of participants with missing data for each variable of interest | 9-10, Table 1, Table 2, Figure 1, Supplementary Table 2 |
| | | (c) Summarise follow-up time (eg, average and total amount) | 9-10, Table 2 |
| Outcome data | 15* | Report numbers of outcome events or summary measures over time | 9-10, Table 2 |
| Main results | 16 | (a) Give unadjusted estimates and, if applicable, confounder-adjusted estimates and their precision (eg, 95% confidence interval). Make clear which confounders were adjusted for and why they were included | 9-10, Table 2 |
| | | (b) Report category boundaries when continuous variables were categorized | Not applicable |
| | | (c) If relevant, consider translating estimates of relative risk into absolute risk for a meaningful time period | 9-10, Table 2 |
| Other analyses | 17 | Report other analyses done—eg analyses of subgroups and interactions, and sensitivity analyses | 10, Figure 2, Figure 3 |
| **Discussion** | | | |
| Key results | 18 | Summarise key results with reference to study objectives | 10-11 |
| Limitations | 19 | Discuss limitations of the study, taking into account sources of potential bias or imprecision. Discuss both direction and magnitude of any potential bias | 12-13 |
| Interpretation | 20 | Give a cautious overall interpretation of results considering objectives, limitations, multiplicity of analyses, results from similar studies, and other relevant evidence | 11-12 |
| Generalisability | 21 | Discuss the generalisability (external validity) of the study results | 12-13 |
| **Other information** | | | |
| Funding | 22 | Give the source of funding and the role of the funders for the present study and, if applicable, for the original study on which the present article is based | 13 |

**Supplementary Table 2.** Associations of characteristics of the schools participating in the School-SARS-CoV-2 Study.

| **No. of schools within categories (column %)** | **School type** | | **Social deprivation index*** | | **Average class size** | |
|---|---|---|---|---|---|---|
| | Primary school | Secondary school | Low/ moderate | High/very high | ≤20 pupils/ class | >20 pupils/ class |
| **Local population density** | | | | | | |
| ≤100 inhabitants / km$^2$ | 17 (13%) | 6 (5%) | 23 (13%) | 0 (0%) | 19 (18%) | 4 (3%) |
| >100-250 inhabitants / km$^2$ | 25 (19%) | 22 (19%) | 47 (26%) | 0 (0%) | 30 (30%) | 17 (12%) |
| >250-500 inhabitants / km$^2$ | 19 (15%) | 15 (13%) | 30 (16%) | 2 (3%) | 21 (21%) | 13 (9%) |
| >500-10000 inhabitants / km$^2$ | 60 (47%) | 66 (57%) | 80 (44%) | 45 (76%) | 31 (31%) | 95 (67%) |
| >10000 inhabitants / km$^2$ | 8 (6%) | 7 (6%) | 3 (2%) | 12 (20%) | 3 (3%) | 12 (9%) |
| | P=0.231 | | P<0.001 | | P<0.001 | |
| **School type** | | | | | | |
| Primary school | | | 93 (51%) | 33 (56%) | 77 (74%) | 52 (37%) |
| Secondary school | | | 90 (49%) | 26 (44%) | 27 (26%) | 89 (63%) |
| | | | P=0.494 | | P<0.001 | |
| **Social deprivation index*** | | | | | | |
| Low/moderate | | | | | 85 (83%) | 98 (70%) |
| High/very high | | | | | 17 (17%) | 42 (30%) |
| | | | | | P=0.017 | |

P values are from $\chi^2$-tests. *Information on the social deprivation index was not available for three schools.

**Supplementary Table 3.** Volumes (in µl) of sample material used for RNA extraction and RT-qPCR across laboratories.

| | Graz | | Innsbruck | Linz | Vienna | |
|---|---|---|---|---|---|---|
| | Round 1 | Round 2 | Round 2 | Round 2 | Round 1 | Round 2 |
| Sample (DTT) | 200 (10) | 200 (10) | 200 (20) | 100 (15) | 100 (12.5) | 100 (12.5) |
| Lysis buffer | N/A | 265 | 200 | 350 | 350 | 350 |
| Elution | N/A | 50 | 100 | 50 | 50 | 50 |
| Template | N/A | 7 | 5 | 7 | 7 | 7 |
| RT-qPCR reaction | N/A | 20 | 20 | 20 | 20 | 20 |
| Viral targets (pool) | E, ORF1a/b | N1, N2, ORF1b, ORF10, RP2 | N2, ORF1b, RP2 | N2, ORF1b, RP2 | N2, UTR, ORF1b, RP2 or N2, ORF1b, RP2 | |
| Viral target (unpooled) | E, ORF1a/b | N2, ORF1b, RP2 | N1, N2, ORF1b, ORF10, RP2 | N1, N2, ORF1b, ORF10, RP2 | N2, ORF1b, RP2 | |

**Supplementary Table 4.** Primers used in this study for detection of SARS-CoV-2 via RT-qPCR.

| Primer name (*gene*) | Target | Type | 5'-MOD | Primer | 3'-MOD | Reference |
|---|---|---|---|---|---|---|
| IMP-RP2 (*RPP30*) | human | Forward | - | AGATTTGGACCTGCGAGCG | - | Lu et al.[1]* |
| | | Reverse | - | GCAACAACTGAATAGCCAAGGT | - | |
| | | Probe | HEX | TTCTGACCTGAAGGCTCTGCGCG | BHQ1 | |
| IMP-ORF1b (*Orf1b*) | viral | Forward | - | TGGGGTTTTACAGGTAACCT | - | Chu et al.[2]† |
| | | Reverse | - | AACACGCTTAACAAAGCACTC | - | |
| | | Probe | TexasRed | TAGTTGTGATGCAATCATGACTAG | BHQ1/2 | |
| CDC-N2 (*N gene*) | viral | Forward | - | TTACAAACATTGGCCGCAAA | - | Lu et al.[1]* |
| | | Reverse | - | GCGCGACATTCCGAAGAA | - | |
| | | Probe | FAM | ACAATTTGCCCCCAGCGCTTCAG | BHQ1 | |
| IMP-UTR (*3'-UTR*) | viral | Forward | - | AGTGTACAGTGAACAATGCT | - | Newly designed |
| | | Reverse | - | ATCACATGGGGATAGCACTA | - | |
| | | Probe | HEX | AGCTGCCTATATGGAAGAGCCCT | BHQ1 | |
| CDC-N1 (*N gene*) | viral | Forward | | GACCCCAAAATCAGCGAAAT | - | Lu et al.[1] |
| | | Reverse | | TCTGGTTACTGCCAGTTGAATCTG | - | |
| | | Probe | FAM | ACCCCGCATTACGTTTGGTGGACC | BHQ1 | |
| IMP-ORF10 (*Orf10*) | viral | Forward | - | TCGCTTTTCCGTTTACGATA | - | Newly designed |
| | | Reverse | - | ACATCTACTTGTGCTATGTAGTT | - | |
| | | Probe | TexasRed | ACTCTTGTGCAGAATGAATTCTCGT | BHQ1 | |

Modifications to published primers: *Alternative reverse primer spanning an exon-junction. †Elimination of ambiguous nucleotides.

#### Supplementary Figures

**Supplementary Figure 1.** Total no. of teachers and pupils at participating schools and no. of teachers and pupils selected to be included in the School-SARS-CoV-2 Study.

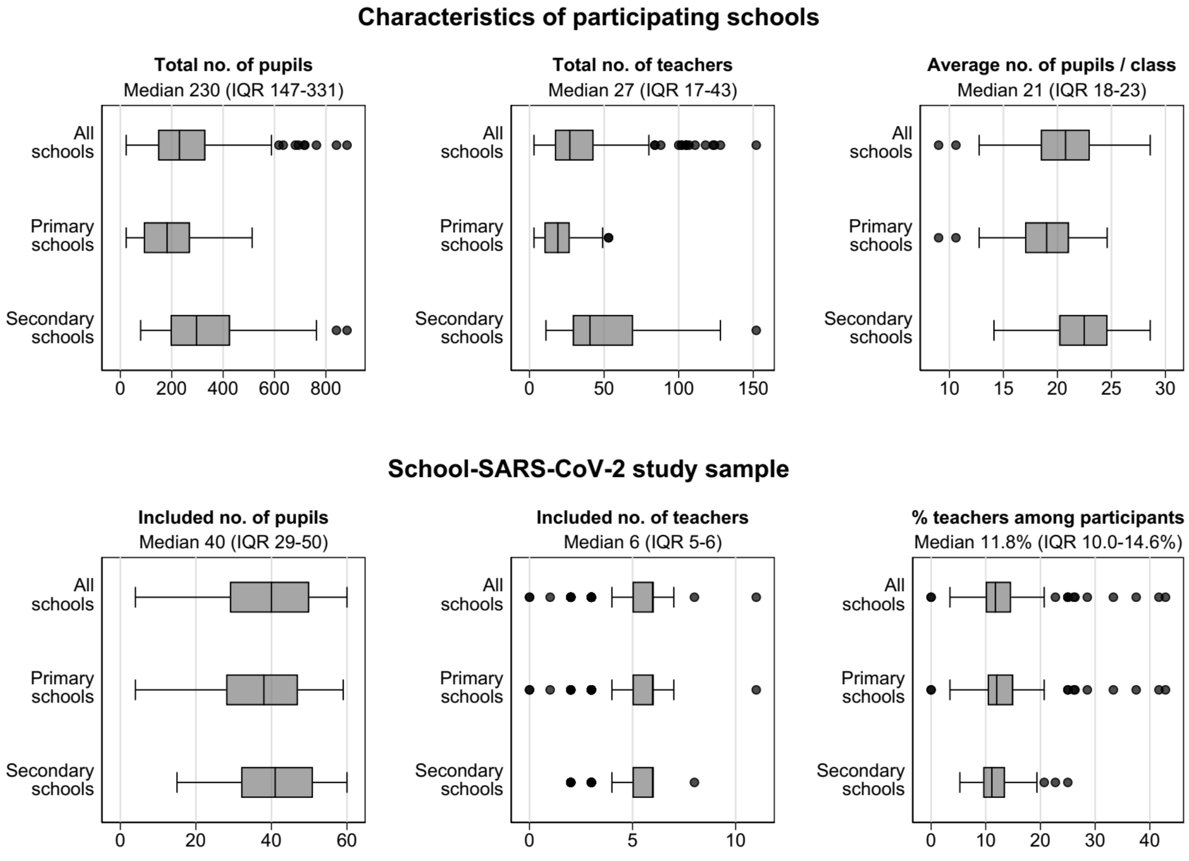

IQR denotes interquartile range. Medians and interquartile ranges included in the figure subtitles refer to the overall study sample (all schools).

**Supplementary Figure 2.** $C_t$ values measured in unpooled samples of positively tested pupils vs. teachers and association with age among pupils.

**(a)** Difference between pupils and teachers

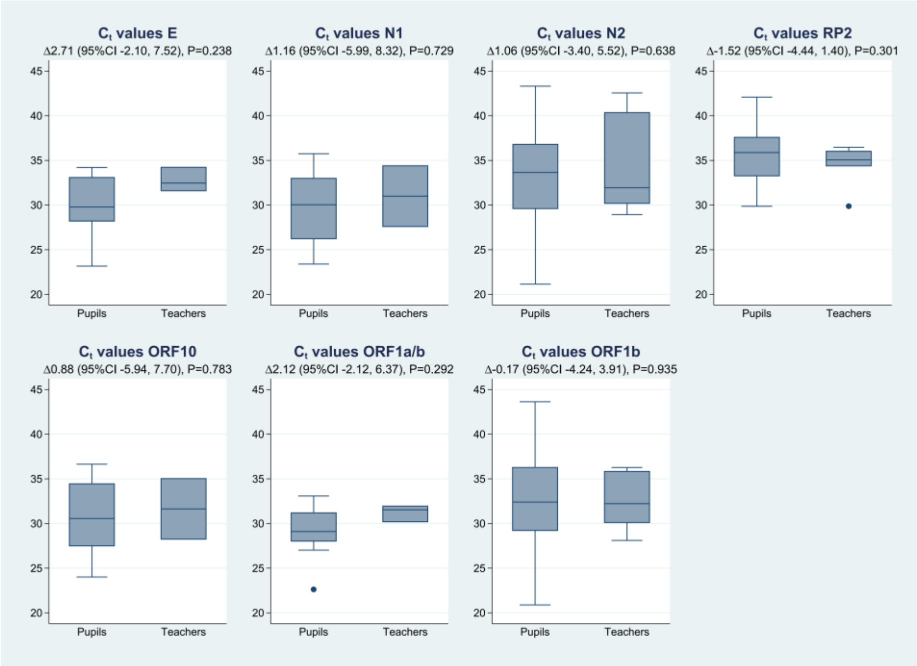

**(b)** Association with age among pupils

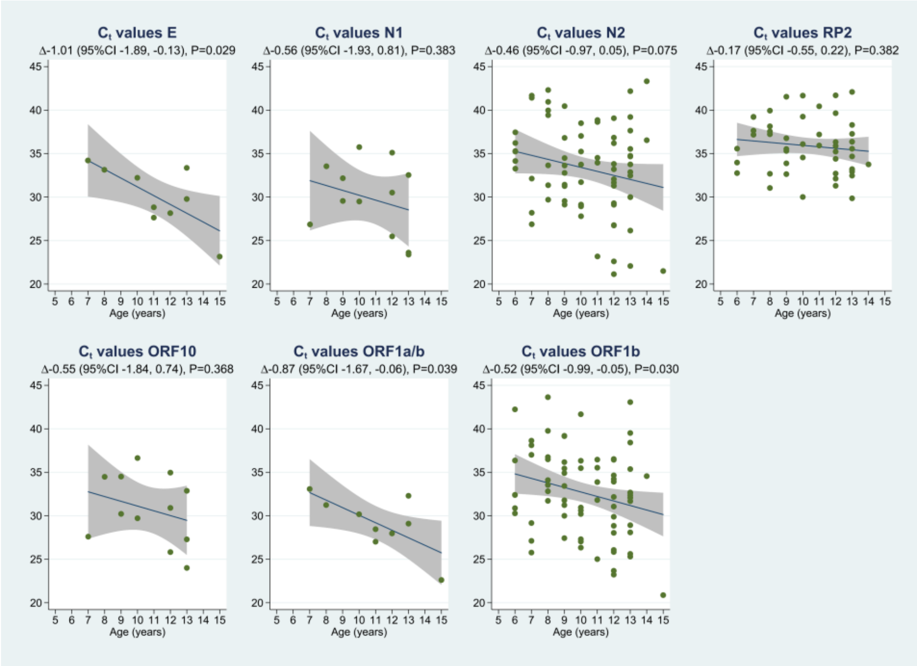

CI denotes confidence interval. The numbers in the subtitles are mean differences in $C_t$ values between pupils and teachers (Panel a) and per one year older age (Panel b) estimated from linear regression models.

**Supplementary Figure 3.** $C_t$ values measured in unpooled samples of participants positively tested for SARS-CoV-2, per testing round and site.

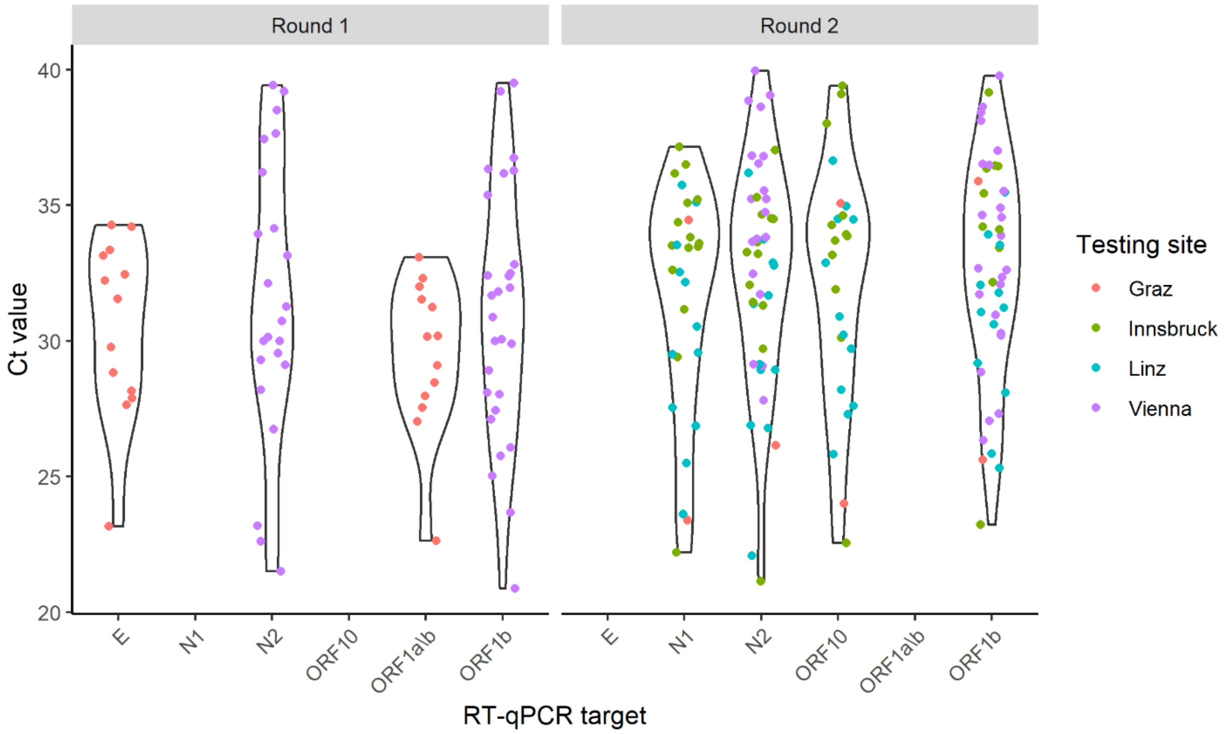

Values >40 are not shown.
